# Investigating CYP2C9*3 Polymorphisms in Patients with Neuropsychological Disorders through Pharmacogenomics Testing

**DOI:** 10.64898/2025.12.03.25340647

**Authors:** Braa Elwaleed, Muhsin Konuk, Tayfun Gözler, Hana Nadir A. Arbab, Abeer Azhary Sayed Abdoon

## Abstract

**Background:** The CYP2C9 gene encodes the cytochrome P450 2C9 enzyme, which plays a key role in metabolizing various drugs. Genetic polymorphisms in CYP2C9 may alter enzyme activity, influencing drug metabolism and treatment response. This study investigates the distribution of CYP2C9*3 polymorphisms and predicted phenotypes in patients with neuropsychological disorders, including bipolar disorder, depression, and schizophrenia, through pharmacogenomics testing.

**Methods:** Blood samples were collected from 20 patients (9 with depression, 6 with bipolar disorder, and 4 with schizophrenia). Genomic DNA was extracted using the Invitrogen CYP2C9 DNA kit. Genotyping was performed with TaqMan Genotyping Assays (Applied Biosystems) on the Thermo Fisher QuantStudio 5 Real-Time PCR system. Allele frequencies were evaluated for the Hardy-Weinberg equilibrium.

**Results:** Among the 20 patients, the distribution of CYP2C9*3 genotypes was 60% (n=12) AA and 40% (n=8) AC. Allele frequencies were 80% (n=32) for the A allele and 20% (n=8) for the C allele. Predicted metabolic phenotypes showed 60% extensive metabolizers (EM) and 40% intermediate metabolizers (IM).

**Conclusions:** Our findings indicate that the *1/*1 genotype appears more often than the *1/*3 genotype in patients with neuropsychological disorders. The identification of intermediate metabolizers highlights the potential of pharmacogenomics testing to guide drug therapy. Given the small sample size, results should be interpreted with caution. Future studies with larger groups, more CYP2C9 variants, and post-treatment follow-up are needed to better understand genetic influences on drug metabolism.

## Introduction

According to the WHO, globally, one in eight people will likely experience mental illness or neurological disorders in their lifetime [1]. Neuropsychological diseases are a group of mental disorders that impair brain function and behavior, significantly disrupting daily life (e.g., schizophrenia, bipolar disorder, depression). Depression involves a sad mood, loss of interest, trouble concentrating, and suicidal thoughts [2,4]. Bipolar disorder can be seen as mania (activity, irritability, insomnia, confidence) or depression (low mood, low energy) [5]. Schizophrenia is tied to dopamine problems, causing positive symptoms (delusions, hallucinations) or negative ones (lack of motivation, flat affect) [6].

These disorders are primarily treated with antipsychotic or antidepressant drugs that modulate neurotransmitters such as dopamine, serotonin, and norepinephrine [7]. However, these treatments often fail for some patients, and many report adverse effects, supporting the use of pharmacogenomic methods for personalized therapy [8]. Additionally, pharmacogenomic tests (PGx) have recently emerged as precision medicine tools that identify genetic variants influencing drug response [9, 10]. Pharmacogenetics, which began in the 1950s, examines how inherited genetics influence drug action and toxicity, primarily through genes related to metabolism [11].

Pharmacogenomic testing uses this data to make drug treatment safer and more effective, which is key to personalized medicine. Cytochrome P450 (CYP450) is a family of enzymes distributed throughout tissues [12]. These enzymes detoxify both endogenous and exogenous substances through multiple metabolic processes. Genetic variation in CYP enzymes results in distinct metabolic patterns, influencing drug metabolism in individuals and informing personalized therapy [12, 16]. CYP enzyme allelic differences result in phenotypes characterized by normal metabolism (NM), intermediate metabolism (IM), rapid metabolism (RM), or ultra-rapid metabolism (UM) [17]. Mutations can change enzyme structure and function [14, 18].

CYP2C9 polymorphisms can affect the binding site [11, 14, 19]. CYP2C9 includes *1, *2, *3, *5, *6, *8, *11, and 13 alleles [20]. CYP2C9*3 is a non-functional allele that reduces drug metabolism [17, 20].

This study examines the impact of the CYP2C9*3 allele in patients with neuropsychological disorders (bipolar disorder, depression, and schizophrenia) using pharmacogenomic analysis, focusing on its influence on medication effectiveness, drug choice, and personalized treatment plans.

## 2. Methodology

### 2.1. Patient Selection

This study is a prospective cross-sectional study conducted at a tertiary psychiatric care center. Patients with neuropsychiatric disorders (depression, bipolar disorder, and schizophrenia) within a broad age range were evaluated and included if they met the DSM-5 diagnostic criteria. Two qualified mental health professionals independently confirmed all diagnoses during routine clinical assessments. No specific dates, locations, or identifiable clinical details were collected or reported.

### 2.2. Sample Collection

A total of 20 whole-blood samples were collected in EDTA tubes from participants diagnosed according to DSM-5 criteria. Demographic variables, including age (reported as non-overlapping 5-year age ranges), gender, and diagnosis, were documented using a standardized questionnaire. All laboratory procedures were performed in an accredited molecular diagnostics facility. Written informed consent was obtained from all participants before inclusion in the study.

### 2.3. DNA Isolation

The Invitrogen CYP2C9 genomic DNA kit was used to obtain DNA from blood samples (Invitrogen, USA). Two hundred microliters of blood were taken, followed by the addition of 200 µL of binding buffer and 20 µL of proteinase and RNase. The mixture was then vortexed. The mixture was incubated for 10 minutes at 55 °C. One hundred microliters of ethanol was added, and the mixture was vortexed again to ensure complete DNA precipitation. The mixture was transferred to a filter collection tube and centrifuged at 11,000 rpm for 3 minutes. Two centrifugation processes were performed: the first with 500 µL of “wash buffer 1” and the second with 500 µL of “wash buffer 2,” both of which were centrifuged at 14,000 rpm for 3 minutes. After that, the filter collection tube was discarded and replaced with a 1.5 mL Eppendorf tube.

Finally, 100 µL of elution buffer was added, and the mixture was centrifuged at 14,000 rpm for 2 minutes. The filter was discarded, and the remaining solution contained DNA.

### 2.4. CYP2C9*3 Gene Polymorphism Genotyping with Thermo Fisher QuantStudio 5 Real-Time PCR Kit

Genotyping analyses were performed using TaqMan Genotyping Assays (Applied Biosystems, Foster City, CA, USA) on the Thermo Fisher QuantStudio 5 Real-Time PCR system (Thermo Scientific, Waltham, Massachusetts, USA). DNA samples were obtained from patients with depression, bipolar disorder, and schizophrenia. Out of the total 20 DNA samples, 9 were from depression patients, 6 from bipolar disorder patients, and 5 from schizophrenia patients. These samples were detected by fluorescence using specific probes that hybridized during the annealing step of the PCR cycle for the amplicon.

### 2.5 Preparation of Samples for Real-Time PCR

For CYP2C9*3 (rs1057910) genotyping, the Thermo Fisher QuantStudio 5 system was used. The PCR mix included master mix, distilled water, TaqMan Genotyping Assays, and DNA. Controls (positive: heterozygous DNA; negative: water) were added. Mixtures were loaded into 96-well plates. The plate was sealed with adhesive foil, and the wells were checked for bubbles and tapped to remove any that remained. After final checks, the PCR program started.

### 2.6. Statistical Analysis

IBM SPSS Statistics for Windows, Version 25.0 (Statistical Package for the Social Sciences, IBM Corp., Armonk, NY, USA) was used for the statistical analysis of the data obtained from the genotyping results. The patients’ sociodemographic findings and genotype results were presented as counts and percentages for categorical data and as mean ± SD and median (min–max) for numerical data. The Mann–Whitney U test, a nonparametric test, was used for age and genotype comparisons. Fisher’s exact test was used to compare categorical variables. A p-value of <0.05 was considered statistically significant.

## 3. Results

Characteristic features of each patient are given in the (Age, Gender, and Diseases) Table 1.

**Table 1:**
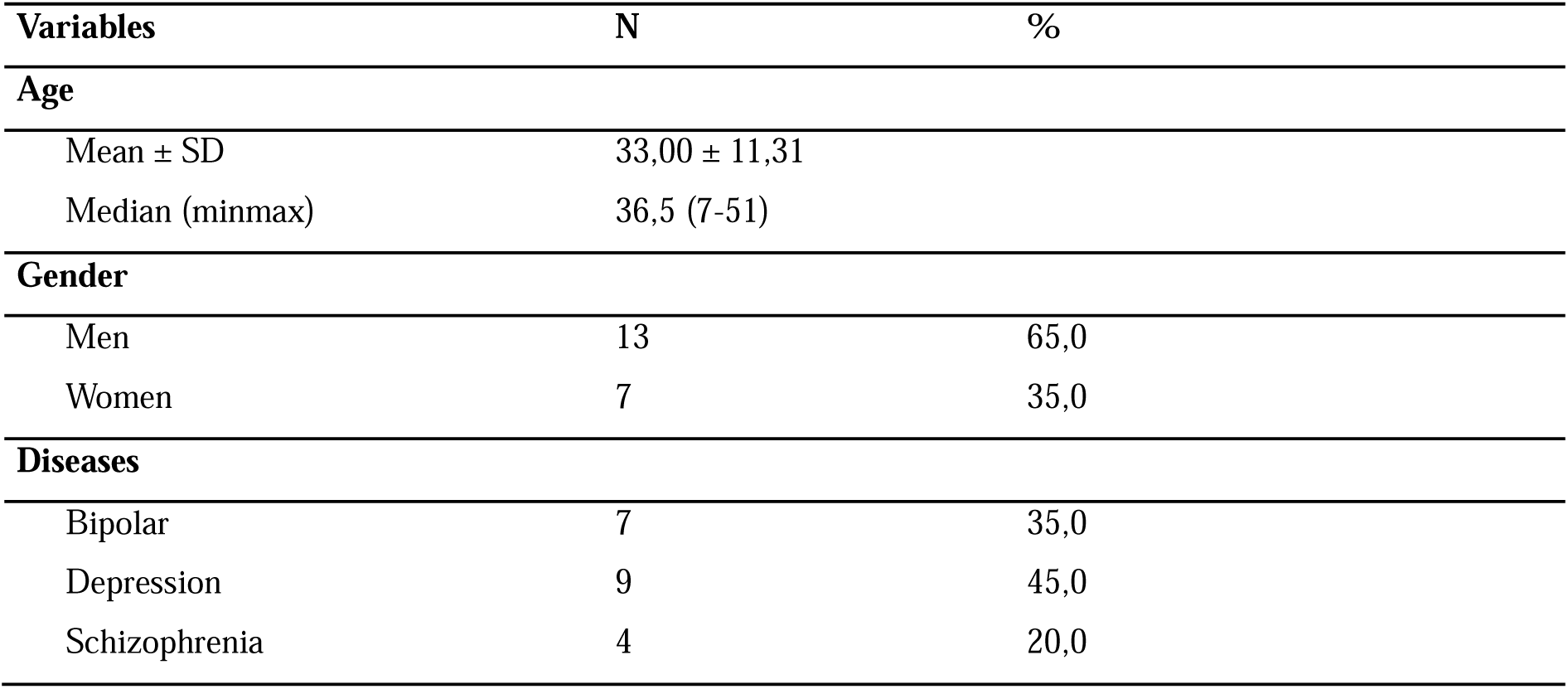
Distribution of character features of patients.

As seen in Table 2, the average age of the patients was 33.00 ± 11.31 years, and the median age was 36.5 (7-51). The minimum age of the patients was 7 and the maximum age was 51. 65% (n = 13) of the patients were male and 35% (n = 7) were female. 35% (n=7) of the patients were diagnosed with bipolar disorder, 45% (n=9) with depression, and 20% (n=4) with schizophrenia.

**Table 2:**
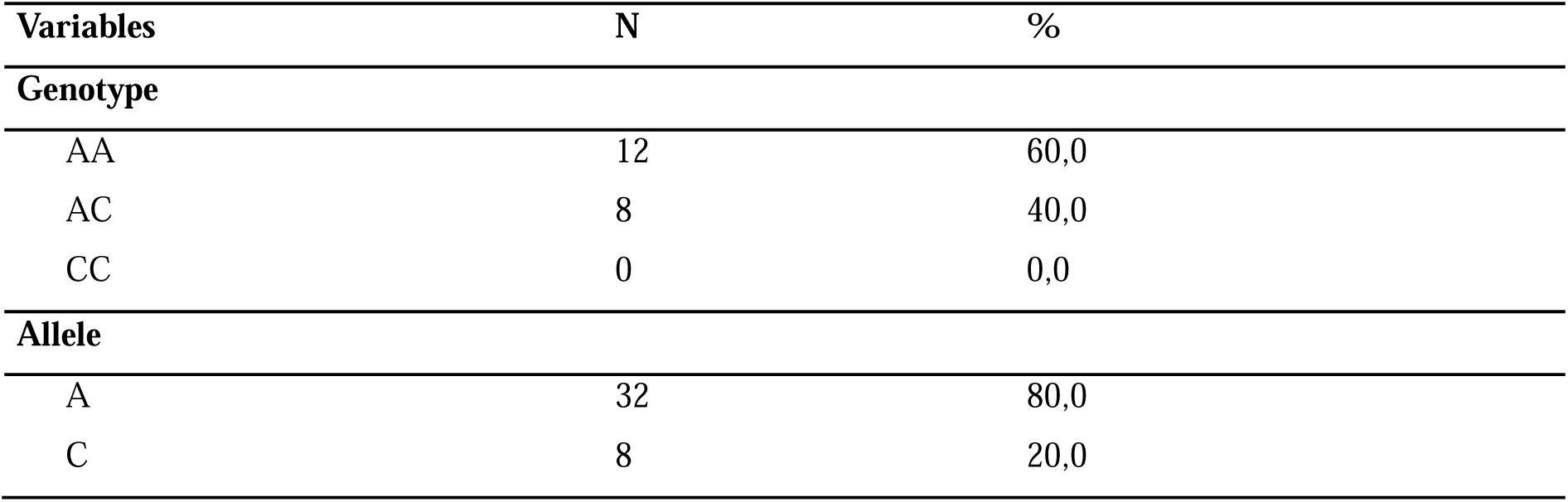
CYP2C9 *3 genotype and allele distributions of patients.

As seen in Table 3, the distribution of the genotypes belonging to the CYP2C9 *3 polymorphism was determined as 60% (n = 12) AA and 40% (n = 8) as AC. When the allele distributions were examined, 80% (n = 32) were determined as the A allele and 20% (n = 8) were determined as the C allele.

**Table 3:**
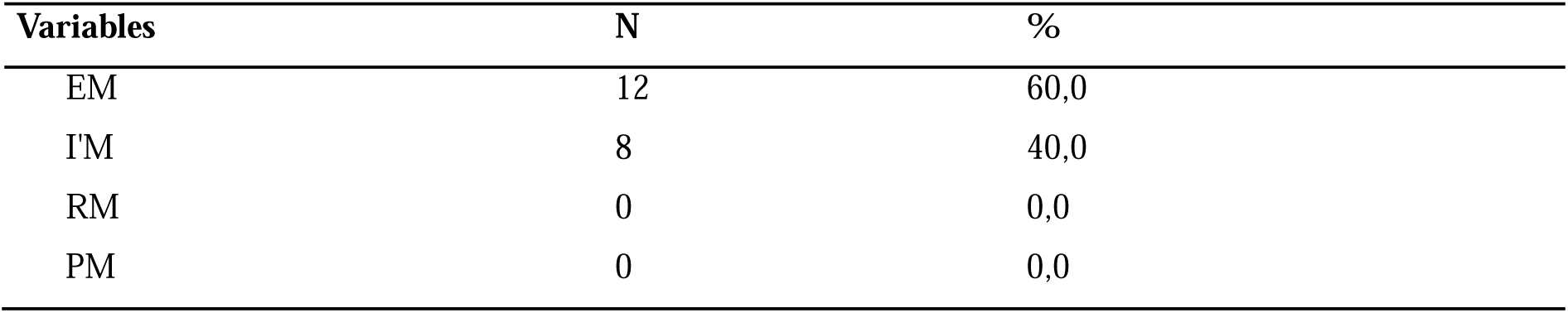
CYP2C9 *3 metabolisms of patients.

As seen in Table 4, the distribution of metabolisms belonging to the CYP2C9 *3 polymorphism was determined as 60% (n = 12) EM-normal metabolism and 40% (n = 8) as IM-Intermediate metabolism.

**Table 4:**
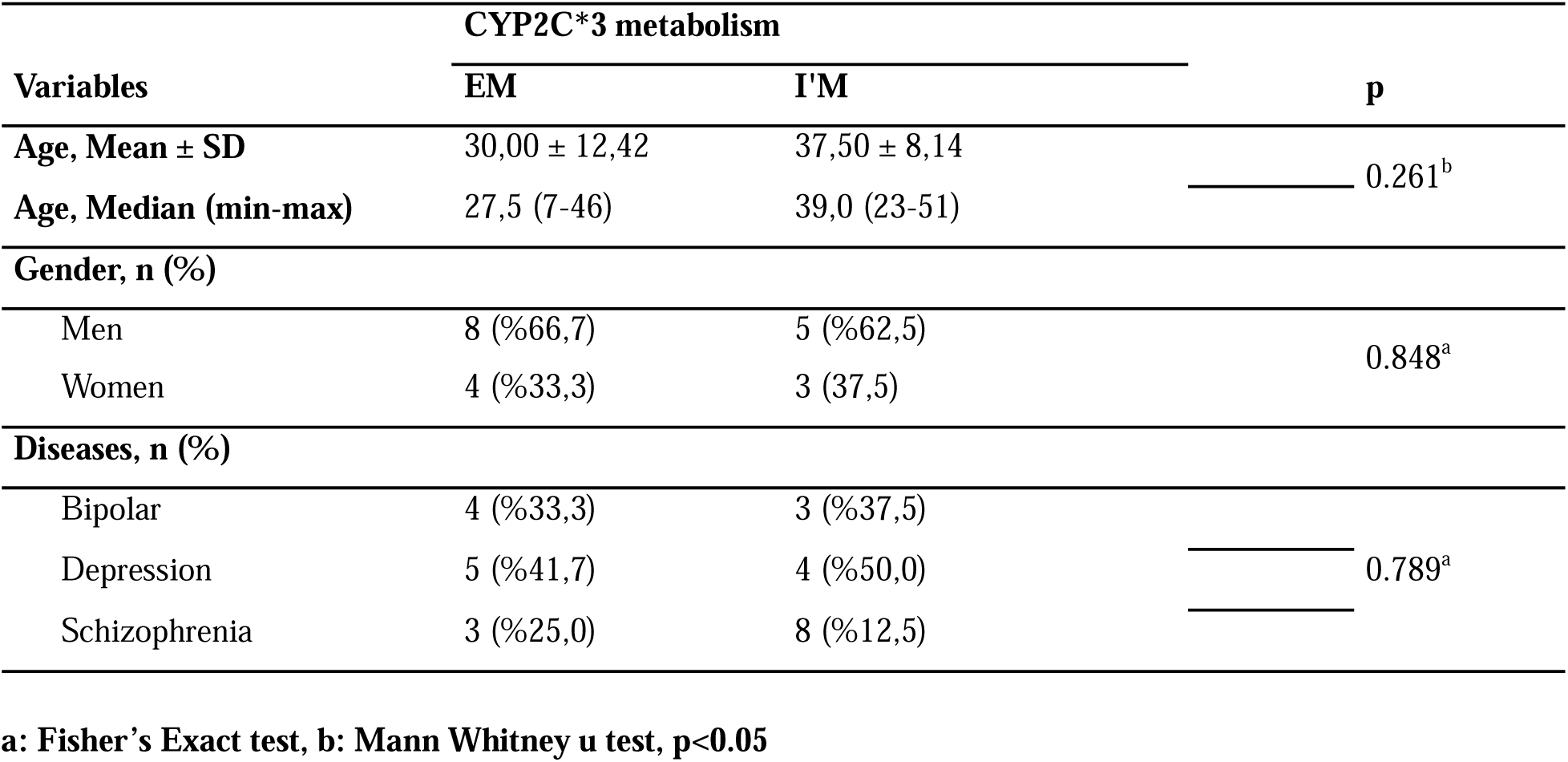
Distribution of characteristic features of patients.

As seen in Table 5, CYP2C9 *3 metabolism groups did not show a significant difference in terms of Age (p = 0.261), Gender (p = 0.848), and disease (p = 0.789).

## 4. Discussion

This study examined 20 patients with neuropsychological disorders (depression, bipolar disorder, and schizophrenia) to assess CYP2C9*3 polymorphism using pharmacogenomics testing. Key findings revealed that 60% of participants possessed the *1/*1 (AA) genotype, while 40% carried the *1/*3 (AC) genotype; no subjects had the *3/*3 (CC) genotype. Thus, the A allele was found in 80% of subjects and the C allele in 20%, clearly indicating the predominance of the AA genotype in this patient sample.

Assessment of metabolic phenotypes showed that 60% of patients were extensive metabolizers (EM) and 40% were intermediate metabolizers (IM). Analysis demonstrated no significant relationship between CYP2C9*3 metabolic phenotype and age, gender, or diagnosis, underscoring that genotype and predicted metabolic activity were consistently distributed across these variables.

When compared with earlier studies across different populations, our results emphasize considerable variability in CYP2C9*3* allele distribution among ethnic groups. For instance, a study of the CYP2C9 allele in 573 individuals from the Korean population showed that only 13 subjects (2.3%) carried the CYP2C93 allele [22], which is substantially lower than the 20% observed in our study. Similar results were obtained in studies of 140 Japanese individuals [23], 394 Chinese individuals [24], and 993 South African individuals [25]. In contrast, a higher prevalence of CYP2C9*3 was identified in a Spanish cohort [26].

The observed CYP2C9*3 frequency in our study population exceeds that reported in several global populations, notably among individuals of Asian and African ancestry. These differences highlight the importance of implementing population-specific pharmacogenomic testing to optimize personalized treatment plans.

In terms of clinical significance, CYP2C9 is most relevant for drugs with a narrow therapeutic index. Reviews suggest that IMs (*1/*3) may need an initial and maintenance dose reduction of 25%, and PMs (*3/*3) a 50% reduction to avoid toxicity [27]. To illustrate, valproate, commonly prescribed as a mood stabilizer in bipolar disorder, is metabolized by CYP2C9. Variants like *1/*3 and particularly *3/*3 have been linked to increased risk of valproate-induced hepatotoxicity, requiring dose reduction or close therapeutic drug monitoring (TDM) [18,28].

Although CYP2C9 is not the main enzyme for most antidepressants and antipsychotics, polypharmacy is a common issue in psychiatric therapy. For patients concurrently taking CYP2C9 substrates (e.g., mood stabilizers or anticonvulsants), the presence of a less-functioning allele can alter the safety and efficacy of treatment. Moreover, evidence from pediatric populations treated with fluoxetine and adult schizophrenia patients on risperidone revealed no significant impact of CYP2C9 variants on treatment outcomes, emphasizing its limited role in SSRI/antipsychotic metabolism [29].

Nevertheless, the relatively high frequency of *1/*3 genotypes across depression, bipolar disorder, and schizophrenia patients in this study suggests that routine pre-treatment PGx screening could enhance medication safety, especially in settings where mood stabilizers or anticonvulsants are used. This aligns with broader trends toward precision psychiatry, where drug–gene interaction awareness is integrated into clinical decision-making to minimize ADRs and maximize therapeutic benefit [12,20].

From a mechanistic perspective, the functional relevance of the *3 allele stems from a single-nucleotide polymorphism (SNP), particularly a 1075A>C substitution. This substitution results in an amino acid change (Ile359Leu) in the coding region of the CYP2C9 enzyme. The resulting structural change impairs the enzyme’s ability to recognize substrates and perform oxidative reactions, resulting in a significant reduction in enzymatic activity [14,20]. As a consequence, individuals with the *1/*3 genotype show slower drug metabolism, increased drug plasma concentrations, and a longer half-life for drugs that are CYP2C9 substrates[17,18]. Understanding this biochemical impact helps explain the pharmacokinetic differences observed clinically in carriers of the *3 allele and supports the rationale for therapeutic drug monitoring in at-risk patients.

This study offers useful insights into the distribution and clinical importance of CYP2C9*3 polymorphisms in patients with neuropsychological disorders, but there are some important limitations. The small sample size reduces the strength and generalizability of the results. Since the study was conducted at a single center, the findings may not apply to other regions or ethnic groups. We focused only on the CYP2C9*3 allele and did not include other significant variants or enzymes, such as CYP2C9*2, CYP2C19, CYP2D6, or CYP3A4, which are important in the metabolism of many psychotropic drugs. This focus narrows the study’s scope and limits the extent to which the results can be applied in clinical decisions. Additionally, we did not collect data on clinical outcomes, such as drug response or side effects, so we cannot directly link genotype to treatment results. Finally, because there were no *3/*3 carriers in our sample, we could not assess poor metabolizer phenotypes, who are often the most at risk.

Future research should address these limitations by studying larger and more diverse groups of people. Including a wider range of genetic tests would help capture the full complexity of pharmacogenetic differences. It is also important to collect data on clinical outcomes, which would support recommendations for adjusting medication based on genotype. Studying the cost and real-world benefits of pharmacogenomic screening before prescribing psychiatric drugs will help guide its use in practice. Using clinical decision support tools within electronic medical records can make genotype-guided prescribing more practical and personalized. These steps would help move psychiatry toward safer and more effective treatments by applying precision medicine.

## Conclusion

This study constitutes a significant contribution to the examination of CYP2C9*3 gene polymorphisms and their predicted phenotypes in individuals with neuropsychological disorders using pharmacogenomic testing. The findings should be interpreted cautiously due to the study’s limitations. Future research should employ larger sample sizes and include additional polymorphisms and enzymes to assess the effects of pharmacogenomic testing on drug response, metabolism, adverse effects, and medication monitoring across neuropsychological and other disorders. Such investigations will inform improved treatment strategies and advance the field of personalized medicine.

## Data Availability

All data produced in the present study are available upon reasonable request to the authors

## Ethics statement

The Üsküdar University Unitential Research of Ethics Assembly 27/03/2020, the meeting of 03 on the date of 03, “Genotypephenotype in neuropsychiatric diseases: Assessment of the Relationship,” decided that this study research project is ethically eligible.

## Declaration of Competing Interest

The authors declare that they have no conflict of interest.

## Funding Statement

No funding was received for this study.

## Notes

### Competing Interest Statement

The authors have declared no competing interest.

### Funding Statement

This study did not receive any funding

### Summary of Updates

This version of the manuscript has been revised to correct the name of the supervising author. No other changes have been made.

